# At-Home Movement State Classification Using Totally Implantable Bidirectional Cortical-Basal Ganglia Neural Interface

**DOI:** 10.1101/2025.02.21.25322475

**Authors:** Rithvik Ramesh, Hamid Fekri Azgomi, Kenneth H. Louie, Jannine P. Balakid, Jacob H. Marks, Doris D. Wang

**Affiliations:** Department of Neurological Surgery, University of California, San Francisco, California, USA

## Abstract

Movement decoding from invasive human recordings typically relies on a distributed system employing advanced machine learning algorithms programmed into an external computer for state classification. These brain-computer interfaces are limited to short-term studies in laboratory settings that may not reflect behavior and neural states in the real world. The development of implantable devices with sensing capabilities is revolutionizing the study and treatment of brain circuits. However, it is unknown whether these devices can decode natural movement state from recorded neural activity or accurately classify states in real-time using onboard algorithms. Here, using a totally implanted bidirectional neurostimulator to perform long-term, at-home recordings from the motor cortex and pallidum of four subjects with Parkinson’s disease, we successfully identified highly sensitive and specific personalized signatures of gait state, as determined by wearable sensors. Additionally, we demonstrated the feasibility of using these neural biomarkers to drive adaptive stimulation with the classifier embedded onboard the neurostimulator. These findings offer a pipeline for ecologically valid movement biomarker identification that can advance therapy across a variety of diseases.

## INTRODUCTION

Movement decoding from invasive human intracranial recordings typically relies on advanced machine learning algorithms programmed into external computers with corresponding outputs for state classification. Despite the significant strides made by brain-computer interfaces (BCIs) in motor and speech restoration after stroke and spinal cord injuries,^1–4^ training data for BCI algorithms are largely collected in laboratory environments, with subjects performing standardized tasks while neural activity is recorded. Implementation of most BCI technology is consequently resource-intensive, time-limited, and unnatural.^5^ Experimental setups are restricted by the rigidity of artificial research-oriented tasks in contrast to the variety and spontaneity of daily activities performed by subjects outside observational encounters. Additionally, subjects are usually tethered to the computer, which limits the ability to study freely mobile movements such as gait.^6^ Together, these practical considerations have limited our understanding of neural activity in the real world.

The recent development of implantable neurostimulators with sensing functions has expanded our knowledge of human brain function and pathophysiology. The advanced capabilities of bidirectional interfaces to simultaneously record neural activity and provide stimulation have enabled investigations of critical brain networks implicated in an array of neurological and psychiatric disorders.^7–17^ These devices have allowed researchers to collect larger datasets across longer timescales to identify intracranial biomarkers of language processing,^18^ seizure risk,^7,8^ pain,^13^ as well as immobile and dyskinetic states in patients with Parkinson’s disease (PD).^14,19^

Despite the ability for bidirectional neurostimulators to identify biomarkers of pathological brain networks, the utility of these devices for classifying movement state in the naturalistic setting is conspicuously unknown. The classification of movement state and type is central to improving the treatment of many circuit pathologies since motor activity affects connectivity within brain networks and modulates the oscillatory patterns of various cortical and subcortical regions.^20,21^ A prime example of this paradigm is found in PD, where medication and movement states alter cortical-basal ganglia network oscillations, although standard treatment with continuous deep brain stimulation (DBS) does not account for these dynamic fluctuations. Current strategies in adaptive DBS (aDBS) have focused on changing stimulation amplitude based on neural biomarkers of low medication state (increased beta frequency [13-30 Hz] synchronization)^22,23^ or dyskinetic episodes (increased narrowband gamma [∼65 Hz] activity),^24^ but not on movement states or specific types of movement. The ability to identify both the occurrence and type of movement in individuals with PD would further enhance aDBS algorithms to target and improve patient mobility.

Specifically, gait disturbances represent one of the most prevalent and debilitating symptoms of PD, making the identification of neural signatures of gait an important first step in designing aDBS protocols to improve gait functions.^25^ A growing body of evidence suggests parkinsonian gait disorders may be better addressed by stimulation frequencies lower than those used for conventional clinical stimulation.^26–30^ However, gait-optimized frequencies are less effective for appendicular symptom control (e.g., tremor, bradykinesia, rigidity).^31,32^ This distinction in therapeutic stimulation settings for appendicular and axial symptoms necessitates reliable and ecological biomarkers of a person’s movement state, which has not been explored in the real world.^23,24,33,34^

To address these areas of need, our study aimed to develop a pipeline for identifying naturalistic neural biomarkers of gait state in patients with PD. By leveraging long-term at-home intracranial recordings paired with kinematic data collection through external wearable sensors, we sought to decode neural signatures of gait using a totally implanted system **(Figure 1)**. To our knowledge, our study represents the first attempt to collect and decode at-home movement state using chronic multisite neural activity in patients with PD. We used wearable sensors (Rover, Sensoplex Inc.) to identify at-home movement state in four patients with PD, while simultaneously recording subcortical local field potential (LFP) and motor cortex electrocorticography (ECoG) signals via an investigational bidirectional neurostimulator (Summit RC+S, Medtronic). By comparing cortical-pallidal oscillatory activity during walking and non-walking epochs, we identified both shared and subject-specific neural oscillatory bands that distinguish these movement states. Using machine learning techniques, we characterized the relative importance of different frequency ranges and brain regions to accurately classify gait state and derived highly sensitive and specific individualized biomarkers of gait. Finally, we tested the accuracy of the on-board classifier to distinguish gait vs non-gait states with these biomarkers using *in silico* simulation. These findings expand our understanding of gait neurophysiology and demonstrate the viability of at-home data collection as a modality for the identification of patient-specific movement state biomarkers for closed-loop aDBS.

**Figure 1.**
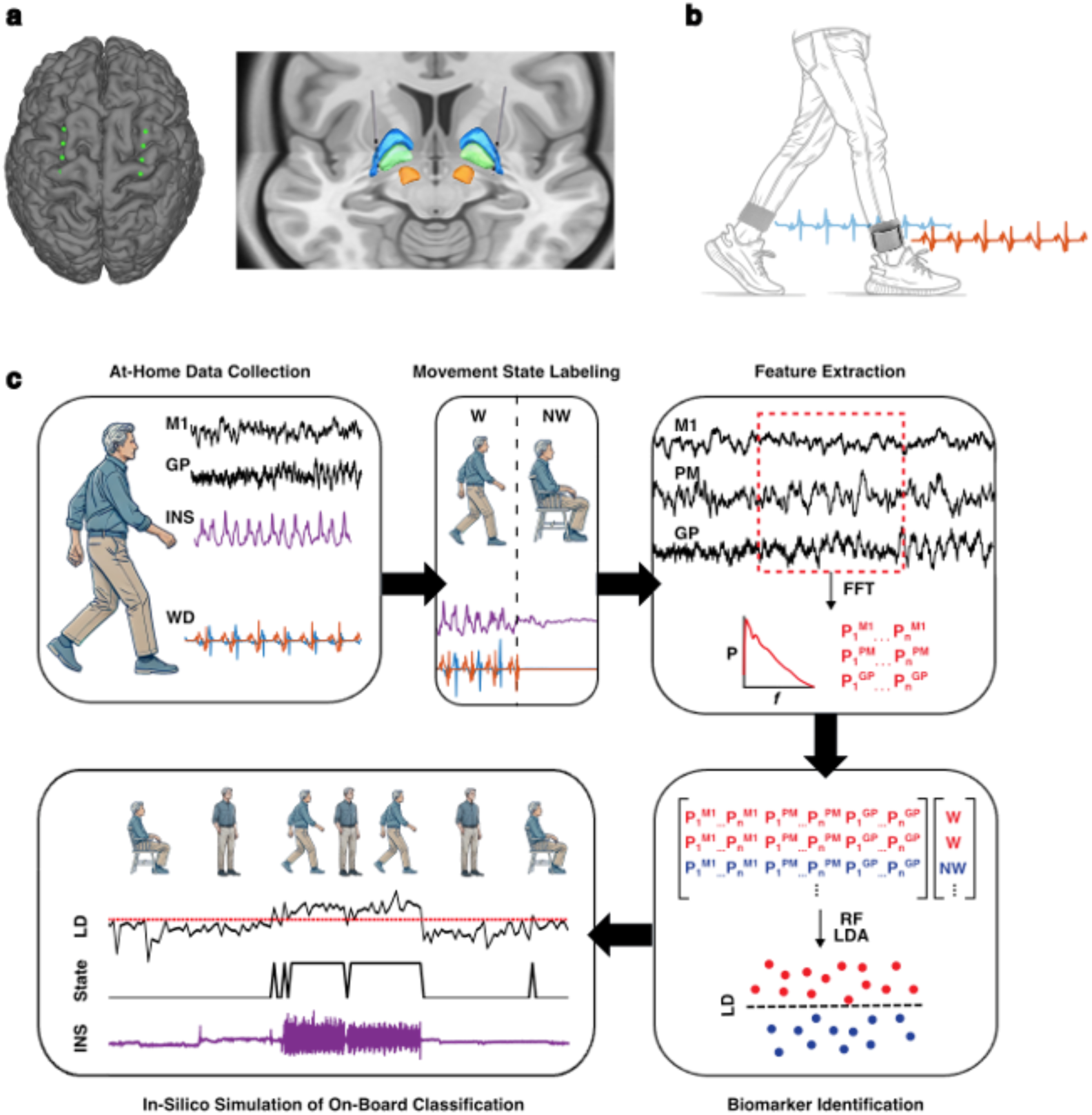
Pipeline for identification of cortical-pallidal neural biomarkers of at-home movement state in Parkinson’s disease. **a.** Example localization of Summit RC+S bidirectional neurostimulator cortical electrodes overlying M1 and PM (left) and subcortical depth electrodes implanted in the GP (right). **b.** Representation of Rover accelerometer WDs, worn around the ankles bilaterally. Sample acceleration signals are shown from the left and right foot. **c.** Schematic representation of neural biomarker identification pipeline. Neural data from M1, PM, and GP and acceleration data from WD and INS were streamed from patients’ homes. Acceleration signals were aligned, partitioned into 10-second epochs, and labeled as periods of continuous walking (W) or non-walking (NW) using WD data. From each epoch, average power was calculated within all possible frequency bands from 1 to 50 Hz. These power value features were used to train and test LDA models. Finally, *in-silico* testing was performed to simulate continuous on-board classification of movement state using system-constrained biomarkers derived from at-home data. FFT = fast Fourier transform; GP = globus pallidus; INS = implantable neurostimulator; LD = linear discriminant; LDA = linear discriminant analysis; M1 = primary motor cortex; NW = non-walking; PM = premotor cortex; RF = random forest; W = walking; WD = wearable device.

## RESULTS

### Patient characteristics and contact localization

Four individuals with PD (2 male, 2 female) undergoing evaluation for DBS implantation were recruited and implanted with a bidirectional investigational neurostimulator device (Summit RC+S, Medtronic). Subject demographics are presented in **Figure 2a**. All participants enrolled in this study exhibited gait dysfunction, which was assessed with the Movement Disorder Society Unified Parkinson’s Disease Rating Scale Part III (MDS-UPDRS III) (score range: 21-57). Specifically, this scale’s posture instability and gait disorder (PIGD) subscore was used to characterize challenges with balance, freezing, and postural instability (score range: 2-10).

**Figure 2.**
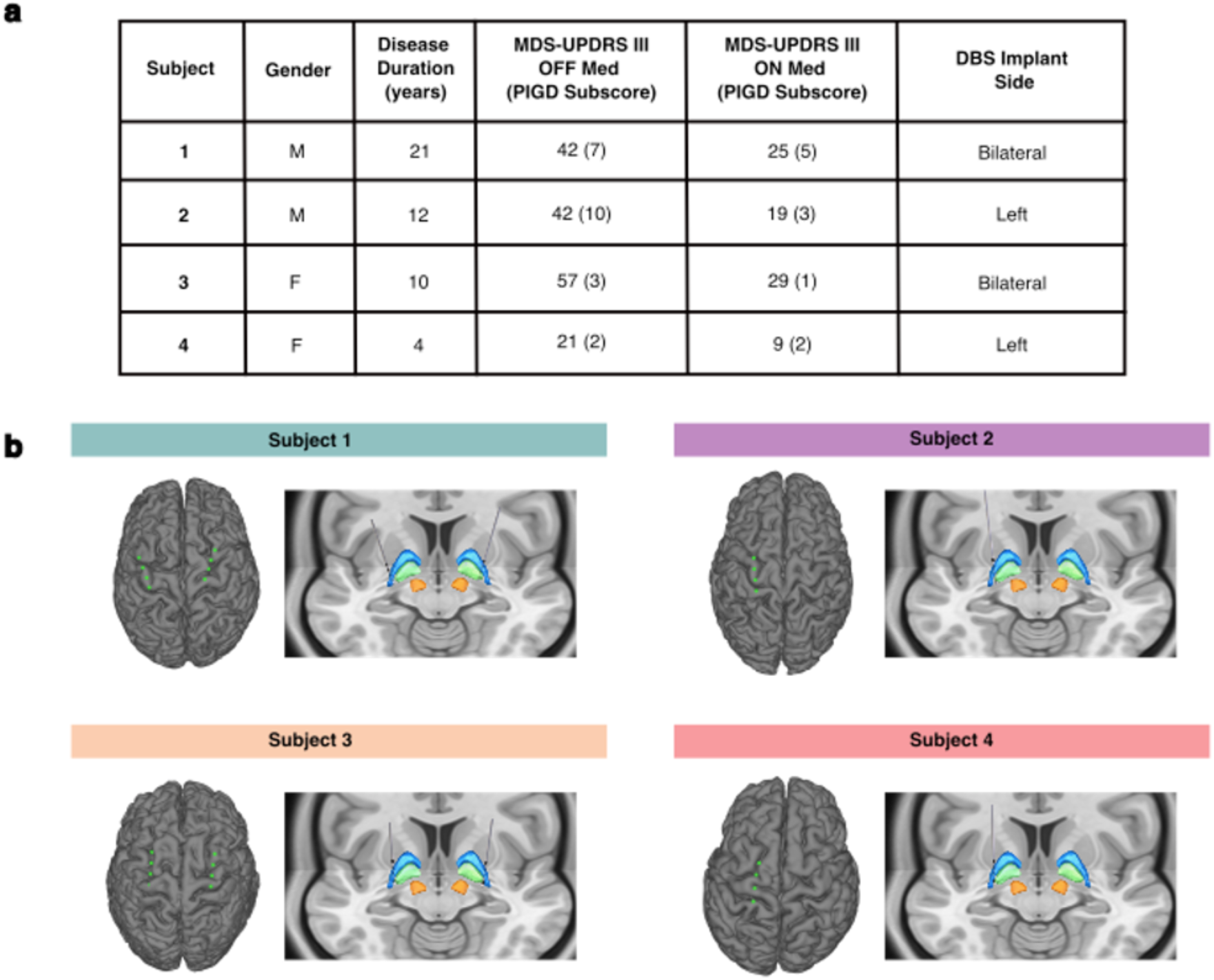
Subject demographics and electrode localization. **a.** Demographic and clinical characteristics are shown for the four subjects enrolled in this study. **b.** Subject-specific reconstructions are shown of ECoG contacts targeting M1 and PM and depth electrodes targeting GP. DBS = deep brain stimulation; ECoG = electrocorticography; GP = globus pallidus; M1 = primary motor cortex; MDS-UPDRS III = Movement Disorder Society – Unified Parkinson’s Disease Rating Scale (Third Revision); PIGD = postural instability and gait disorder; PM = premotor cortex.

Two subjects received unilateral implants (left hemisphere), while the remaining two received bilateral implants. Each DBS device consisted of quadripolar depth leads implanted in the globus pallidus (GP) and subdural quadripolar paddles overlying the sensorimotor cortices. Cortical electrodes recorded neural data from the primary motor (M1) and premotor (PM) cortices in all subjects. Localization of depth and surface electrode contacts is shown in **Figure 2b**. Electrodes were connected to implantable neurostimulators (INS) which generated electrical impulses and streamed kinematic and neural data (Methods).

### Movement state labeling using wearable accelerometer devices

To collect chronic recordings of at-home movement, subjects were provided wearable ankle accelerometer devices (WD) (Rover, Sensoplex Inc), each containing a sensor comprised of a triaxial gyroscope, accelerometer, and magnetometer which recorded kinematic data locally. To validate the ability for these devices to accurately identify walking in our subjects, we collected data from trials of observed walking and non-walking periods in controlled environments. During these in-laboratory trials, data were recorded from participants’ INS, WDs, and force-sensitive resistors (FSRs) placed under the feet bilaterally. Signals from these three devices were aligned and WD-based labeling of movement state was compared with ground-truth labels (Methods). An example data collection session is illustrated in **Figure 3a**.

**Figure 3.**
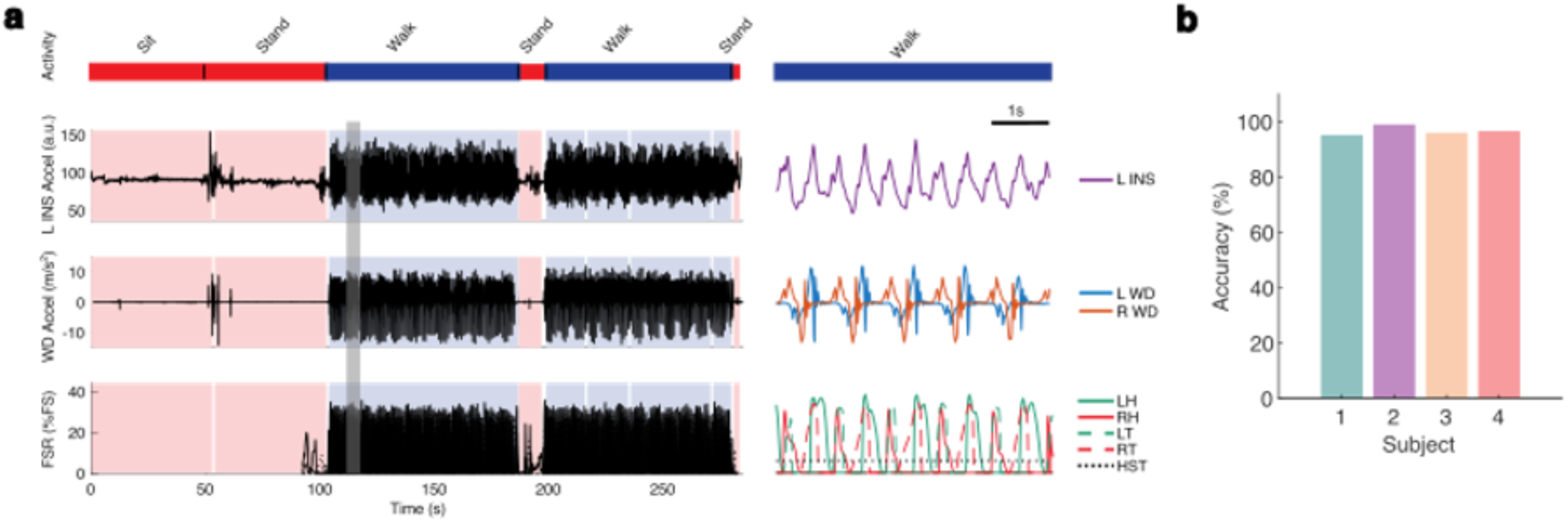
Validation of wearable device (WD) accuracy in labeling walking and non-walking epochs. **a.** Acceleration signals from L INS and bilateral WD are shown aligned with force signals from FSRs on the patient’s feet during a 300 second session of overground walking with interspersed periods of standing or seated rest. The bar at the top indicates patient behavior throughout the session (blue: walking; red: non-walking). A 5 second period of continuous walking indicated by the gray box is enlarged and displayed within the subpanel on the right. **b.** Subject-specific accuracies of WD-based movement state labeling are shown (Supplementary Figure S1b). FSR = force-sensitive resistor; HST = heel strike threshold; INS = implantable neurostimulator; LH = left heel; LT = left toe; RH = right heel; RT = right toe; WD = wearable device.

For all subjects, WDs enabled excellent discrimination of walking and non-walking states **(Figure 3b)**. Accuracy for WD-based labeling was above 95.0% for all subjects, with a range from 95.8% (Subject 1) to 99.0% (Subject 2). Sensitivity varied from 94.4% (Subject 2) to 98.9% (Subject 1) and specificity ranged from 94.7% (Subject 1) to 100.0% (Subjects 2 and 4). See **Supplementary Figure S1** for further detail. These results demonstrate robust WD-based movement state identification across the variety of gait patterns displayed in our subjects and validate the use of WDs to accurately identify periods of walking and non-walking in our cohort **(Supplementary Table S1)**.

### Chronic at-home streaming of kinematic and neural data

Subjects streamed chronic at-home neural and kinematic data from their RC+S INS and WDs as continuously and for as many days as possible. All subjects were receiving clinically optimized continuous GP stimulation during these recordings. Acceleration signals from the INS and WDs were aligned to synchronize neural time-domain data with kinematic data. A sample aligned at-home recording is presented in **Figure 3a**. To facilitate subsequent analyses, neural signals were partitioned into 10-second epochs and WDs were used to label each epoch’s gait state. Across all six hemispheres, a total of 84.5 hours of neural and kinematic data were aligned and analyzed (range: 8.3 to 25.5 hours) from an average of 13 days per hemisphere (range: 5 to 23 days) **(Supplementary Table S2)**.

### Spectral analysis of walking and non-walking epochs

To examine differences in cortical-pallidal neural activity between movement states, we performed within-subject spectral analysis comparing frequency representations between walking and non-walking periods. Power spectral densities (PSD) for each subject and region are presented in **Figure 4b** and **Supplementary Figure S3.** For each 10-second epoch, we computed average power within canonical frequency bands (i.e., delta [1-4 Hz], theta [4-8 Hz], alpha [8-13 Hz], beta [13-30 Hz], and low gamma [30-50 Hz]) for the GP, M1, and PM neural signals.

**Figure 4.**
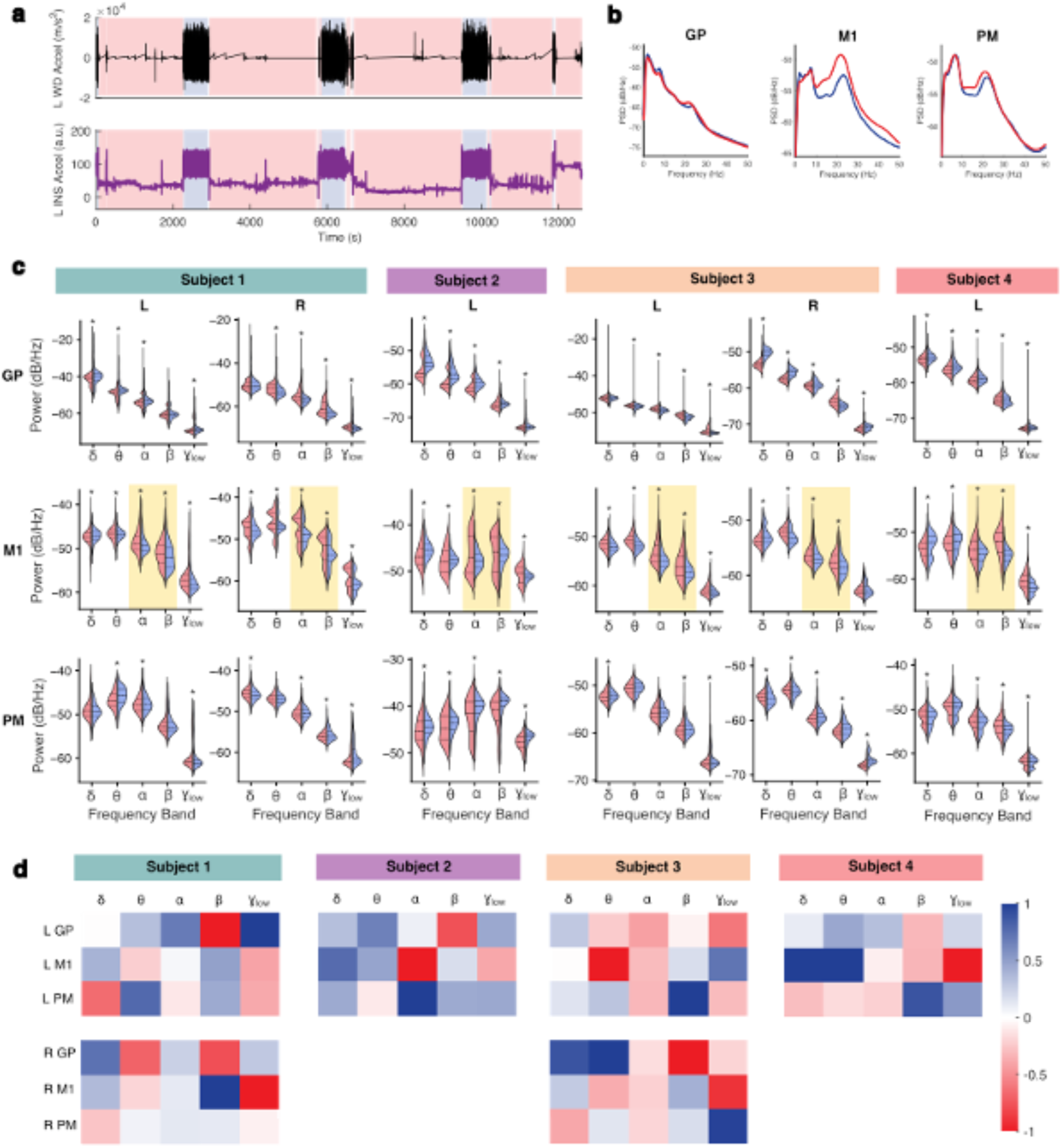
Spectral analysis of at-home walking and non-walking epochs. **a.** Sample at-home recording is shown from Subject 4, with aligned acceleration signals from left WD (top) and INS (bottom). Periods of walking (blue) and non-walking (red) identified with WD-based labeling are indicated with shading. **b.** Sample mean PSDs are shown from 0 to 50 Hz for all walking (blue) and non-walking (red) 10-second epochs analyzed from GP, M1, and PM for Subject 4. **c.** Violin plots are shown comparing mean power within each canonical frequency band between all 10-second non-walking (red) and walking (blue) epochs. Two-sided Wilcoxon rank sum tests were used, with Benjamini-Hochberg correction for multiple comparisons. Across all hemispheres, M1 ɑ and β power was significantly lower during walking epochs, highlighted with yellow boxes. * p < 0.05 (Supplementary Table S4). **d.** Normalized mean feature coefficients from logistic regression classifiers of movement state are visualized for all hemispheres. δ = delta (1-4 Hz); θ = theta (4-8 Hz); ɑ = alpha (8-13 Hz); β = beta (13-30 Hz); γ_low_ = low gamma (30-50 Hz); GP = globus pallidus; INS = implantable neurostimulator; M1 = primary motor cortex; PM = premotor cortex; WD = wearable device.

Overall, when comparing canonical frequency band power across movement states, we observed significantly lower M1 alpha and beta power during walking compared to rest in all subjects (two-sided Wilcoxon rank-sum test; p ≤ 10^-4^) **(Figure 4c)**. Additionally, M1 low gamma power was lower during walking in all but Subject 3’s right hemisphere (p ≤ 10^-9^). Trends in power within all other canonical bands and regions differed by individual; summary power values and p-values are detailed for all comparisons in **Supplementary Table S5**.

In Subject 1, left GP power was higher across all canonical frequency bands except beta during walking (p ≤ 10^-19^) while right GP power was lower in all bands except delta (p ≤ 10^-21^). With the exception of delta power in the left M1, bilateral M1 power was decreased across all bands during walking (p ≤ 0.033). PM power exhibited relative variability bilaterally.

Subject 2 demonstrated significantly increased GP and PM power in all five canonical frequency bands during walking epochs compared to non-walking epochs (p ≤ 10^-14^). In M1, walking was associated with higher low-frequency (delta and theta) power and lower high-frequency (alpha, beta, and low gamma) power (p ≤ 10^-5^).

In Subject 3, left GP power was lower in all canonical bands except delta during walking epochs (p ≤ 10^-15^) while right GP power was higher in all bands except beta (p ≤ 0.001). Left M1 power was decreased in all bands during walking epochs (p ≤ 10^-8^), while right M1 power was more variable. Similar to Subject 1, PM power showed bilateral variability.

In Subject 4, GP power in delta, theta, alpha, and low gamma bands was higher during walking while beta power was lower (p ≤ 10^-27^). In M1, walking epochs were associated with higher delta and theta power, and lower alpha, beta, and low gamma power (p ≤ 10^-5^). In PM, power within all frequency bands except theta was lower during walking (p ≤ 0.003).

To assess the relative contribution of each frequency band to movement state classification, we constructed subject- and hemisphere-specific logistic regression models to classify walking vs non-walking states. Significant decoding performance was achieved in all four subjects (one-sided empiric permutation-based p-values < 0.001) **(Supplementary Table S4)**. We visualized mean regression coefficients rescaled from - 1 to 1 for each subject to identify the most important frequency bands across individuals **(Figure 4d)**. Normalized coefficients and further model details are presented in **Supplementary Table S4**. This analysis revealed that both the most influential canonical power band and most important region varied widely between participants and even between hemispheres in subjects with bilateral implants. These findings suggest that the oscillatory markers of movement state differ between individuals.

### Identification of personalized cortical-pallidal movement state biomarkers

While canonical frequency bands offer the advantage of standardized comparison across individuals, they also carry several limitations including the potential masking of endogenous narrowband peaks.^35^ It is possible that there exist subject-specific frequency bands that enable more accurate differentiation of walking and non-walking than these generalized canonical ranges. To account for this important consideration, we shifted our focus from canonical ranges to all possible frequency bands of sizes 1 to 49 Hz in the range of study (1-50 Hz), which was selected to minimize contributions from clinical stimulation frequencies and subharmonics.

First, using random forest (RF) models, we calculated the importance of each frequency band to accurate discrimination of gait state. By visualizing these feature importances rescaled from 0 to 1 for each subject, we found notable variety in both the regions and bands that supported classification for each subject and hemisphere **(Figure 5a)**. The most important bands for each subject and region are presented in **Supplementary Table S5**. Pallidal features were the most important for most models (4/6), tending to focus on bands falling within delta, theta, and beta range ranges. Notably, in the hemispheres where delta and theta GP activity demonstrated a peak in feature importance, these ranges showed higher activity during periods of walking (Subject 1 left, Subject 2, and Subject 3 right). Contrarily, in hemispheres where beta range GP activity demonstrated a peak in feature importance, pallidal beta activity was lower during periods of walking (Subject 1 right, Subject 3 left and right, Subject 4) or, in the remaining case, not significantly different between movement states (Subject 1 left). Pallidal features were followed in importance by those from M1, which were similarly distributed in low delta and theta frequencies or captured the mid to high beta range. PM features were consistently the least important; on average, the most important features within this region reflected higher frequency bands than those in GP and M1.

**Figure 5.**
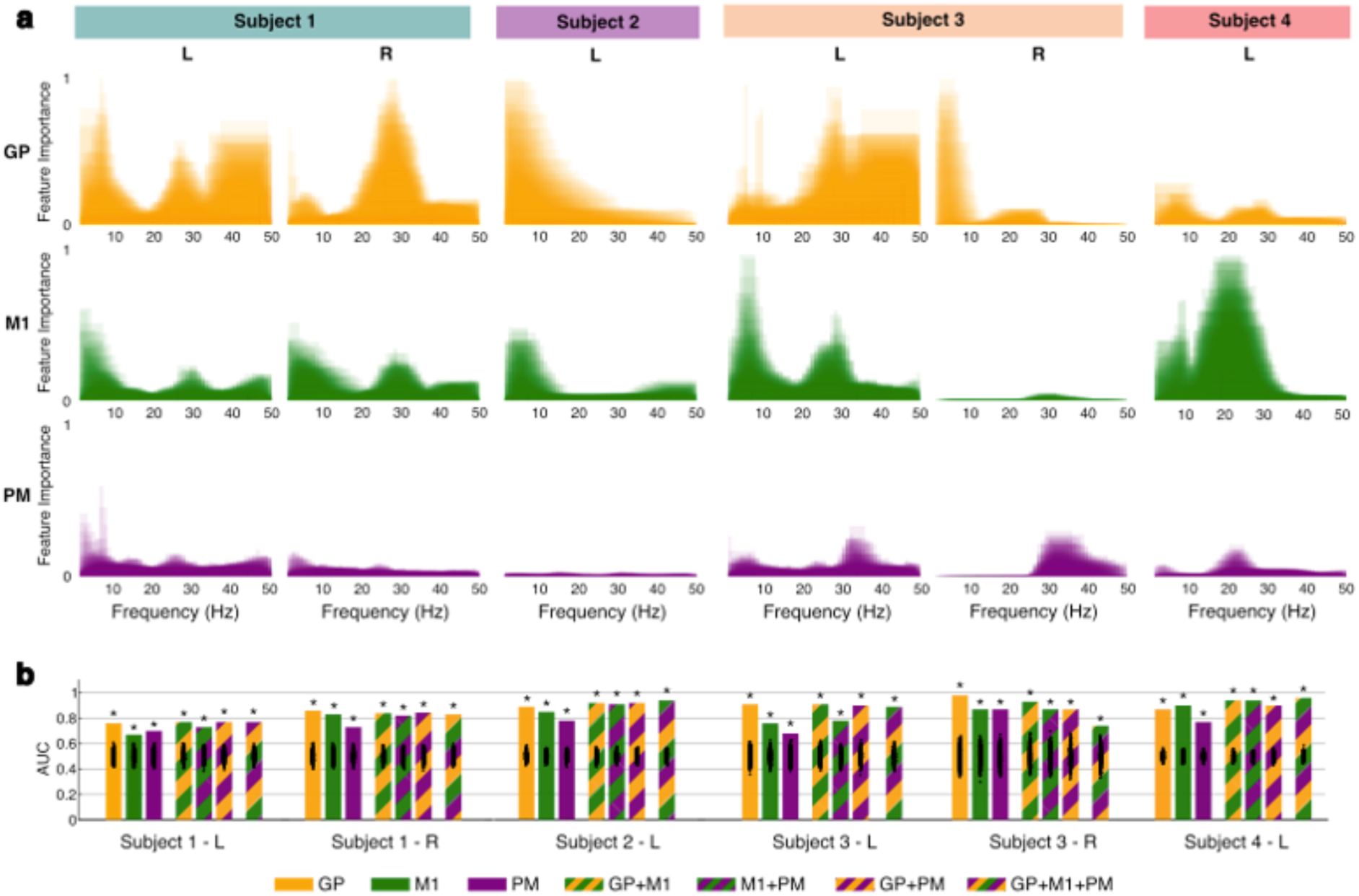
Identification of subject-specific movement biomarkers. **a.** Normalized feature importance is shown for all cortical-pallidal frequency bands using mean MDI from 1,000 RF iterations. **b.** LDA movement state classifier performance is shown for each hemisphere and type of model (single-region, multi-region, and complete). Black dots indicate permuted chance-level performance (n = 1,000), which were used to calculate one-sided empirical p-values. * p < 0.001 (Supplementary Table S6). GP = globus pallidus; LDA = linear discriminant analysis; M1 = primary motor cortex; MDI = mean decrease in impurity; PM = premotor cortex; RF = random forest.

We next trained and tested 10-fold cross-validated LDA models to classify movement state using all frequency band features. Separate models were constructed for each hemisphere using features from all combinations of brain regions: single-region (GP, M1, PM), multi-region (GP+M1, M1+PM, GP+PM), and complete (GP+M1+PM). In all hemispheres, successful movement state classification was possible with activity from a single region (AUC range: 0.67-0.98, p < 0.001) **(Figure 5b** and **Supplementary Table S6)**. Models using pallidal data were the best-performing single-region models in all individuals except for Subject 4, in whom the M1 model was superior. In fact, in three of the six independent hemispheres analyzed, the GP single-region model led to the highest binary classification AUC of any type of model tested (AUC range: 0.86-0.98, p < 0.001). In the remaining cases, the best-performing models were either multi-region or complete models incorporating data from the GP (AUC range: 0.77-0.96, p < 0.001). The best-performing models across all subjects ranged in sensitivity from 74.8-95.8% and specificity from 66.9%-94.0%. Positive predictive value (PPV) was similarly high, varying from 69.7-94.1%. These results support the hypothesis that cortical-pallidal oscillatory activity is modulated by movement state.

### Simulation of on-board movement state classification

The use of neural biomarkers to drive closed-loop aDBS is limited by the technical requirements of the classifiers on-board adaptive-enabled neurostimulators. Therefore, to evaluate the ability for at-home movement state biomarkers to direct actual closed-loop aDBS in practice, we performed in-silico simulation of movement state classification on-board the Summit RC+S device.

First, we simulated classification of at-home movement state using models trained on at-home data. Classification using one to four features meeting on-board constraints was tested (see Methods for further detail). Above-chance performance was achieved for at least one model in all hemispheres (p-value range: <0.001 to 0.006) **(Figure 6a** and **Supplementary Table S7)**. AUC for the best-performing simulations ranged from 0.64 (Subject 1 left) to 0.81 (Subject 4) (p < 0.001). In all hemispheres, the best-performing models were those using three or all four of the permissible features. These models varied in sensitivity (66.0-83.3%), specificity (51.6-69.1%), and PPV (59.5-71.3%).

**Figure 6.**
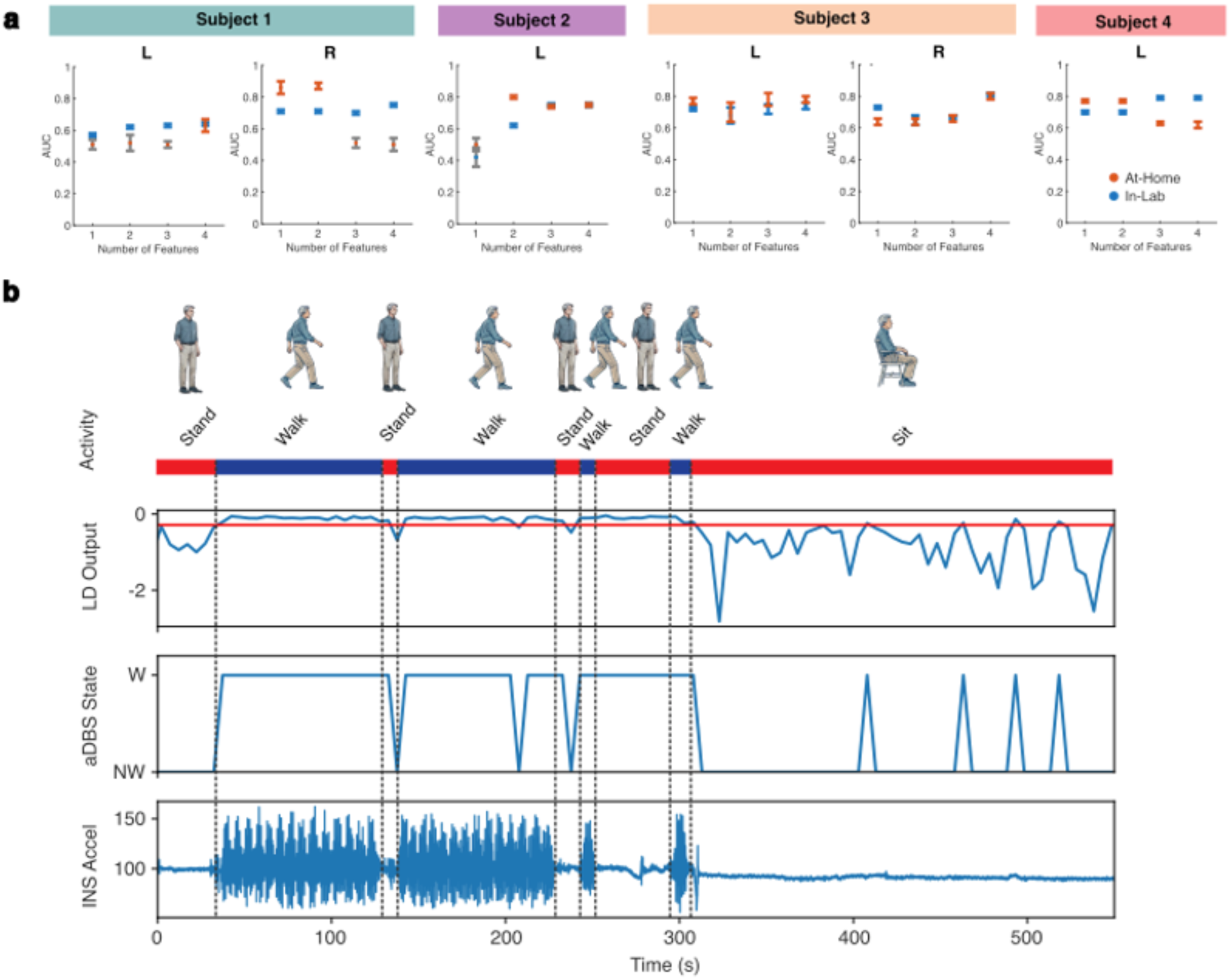
In-silico simulation of on-board movement state classification. **a.** Performance of simulated on-board LDA classifiers is shown for models tested on at-home data (blue) and validated with testing on in-lab observed trials of overground walking with interspersed periods of standing or seated rest (orange). Empirical one-sided p-values were calculated by comparing model performance with a permuted chance-level distribution (n = 1,000) (Supplementary Table S7). Models deemed statistically insignificant (p ≥ 0.05) are shown with grayed bars. **b.** Sample in-silico simulation of continuous on-board classification using 5-second epochs is shown for Subject 1 using biomarker derived from at-home data. An observed trial is shown with true movement state indicated by the bar at the top. Simulated LD output is shown with the threshold indicated by red line, followed by the corresponding aDBS state and true INS acceleration signal. aDBS = adaptive deep brain stimulation; LD = linear discriminant; NW = non-walking; W = walking.

We next validated decoding performance by training simulated on-board classifiers with at-home data and testing these models on in-lab observed trials of overground walking with interspersed periods of standing or seated rest. Again, above-chance performance was demonstrated in all hemispheres, with best-performing models’ AUC varying between 0.63 (Subject 1’s left hemisphere) and 0.85 (Subject 1’s right hemisphere) (p < 0.001) **(Figure 6a** and **Supplementary Table S7)**. Sensitivity (range: 73.0-90.2%), specificity (range: 59.1-81.3%), and PPV (range: 61.5-82.8%) similarly differed across models. Unlike simulations of at-home movement state classification, the best-performing models were not consistently those that used three or four features. A sample on-board in silico simulation of continuous movement state classification is visualized for Subject 1’s right hemisphere in **Figure 6b**. These findings serve as a proof-of-concept demonstration of the feasibility of using at-home data to derive effective movement state biomarkers for closed-loop adaptive stimulation.

## DISCUSSION

Sensing-enabled interfaces now offer the opportunity to dynamically modulate stimulation in response to changes in brain activity, though a major challenge has been the study of complex behavior and identification of ecologically valid neural biomarkers. In particular, the ability to accurately discern naturalistic movement states represents a critical gap in designing neuromodulation therapies targeting specific symptoms. Here, using wearable devices and a novel totally implanted bidirectional neurostimulator, we provide the first in-human evidence for classification of a specific movement type at home from over 80 hours of synchronized kinematic and neural data. Comparing frequency representations within cortical-pallidal signals, we discovered shared and subject-specific spectral features enabling accurate discrimination of walking and non-walking periods. With machine learning techniques, we characterized the variable importance of canonical and non-canonical frequency bands and developed highly sensitive and specific individualized cortical-pallidal signatures of gait. Finally, we constrained biomarkers to the practical limitations of the classifier embedded on-board the Summit RC+S neurostimulator and tested their performance, showing above-chance decoding in all hemispheres. These results have important implications for our understanding of the supraspinal oscillatory dynamics of locomotion and demonstrate the feasibility of successfully collecting and decoding movement state using chronic at-home multisite neural recordings in patients with PD.

Our pipeline provides an effective approach for discovering brain circuit control of natural behaviors in freely-moving humans, which have proved difficult to study in the laboratory setting. Specifically, our understanding of human gait has been limited by methodological constraints. Studies of supraspinal control of gait are typically performed with EEG systems, externalized deep brain stimulation leads, in operating rooms and laboratories. As gait is a highly dynamic state that requires constant adapting and updating based on environmental changes and behavioral goals, studying this movement in an artificial environment presents a significant challenge. However, understanding how gait-related circuits are modulated in the real world with changes in the surrounding environment is critical for treating dynamic symptoms such as freezing of gait, which are often triggered by directional changes, crowded spaces, or emotional states which cannot be easily replicated in experimental settings.^36^

Our approach of identifying neural signatures of gait across different medication cycles over long-term recordings at home yielded important insights about circuit dynamics during walking. Notably, we found lower M1 alpha and beta power and higher GP low-frequency delta and theta activity during periods of walking compared to rest. Oscillatory patterns reflect synchronized activity across neural ensembles and may represent modulations in neuronal excitability that enable the encoding and processing of information within brain networks.^37–39^ Studies of human bipedal walking have demonstrated gait cycle-related changes in brain rhythms, though few have utilized invasive methods of recording neural activity and even fewer have recorded data outside laboratory conditions.^40–44^ In our study of freely ambulating subjects, lower alpha and beta power in M1 during gait align with prior demonstrations of alternating suppression of these bands during periods of walking.^45^ One potential explanation is that desynchronization within these frequency ranges may act to dampen irrelevant sensorimotor signals and direct the spatiotemporal precision of motor activity.^21,46–48^ Our findings also showed that a single region was sufficient to classify movement state in all hemispheres; the preferential importance of GP features to gait decoding seen in most cases supports the hypothesis that pallidal oscillatory activity is central to movement. The most important features from GP signals tended to be narrowband activity from the delta, theta, and beta ranges. Low frequency (delta, theta) GP activity has previously been associated with dystonia, suggesting that synchronization in these ranges promotes a prokinetic state, perhaps signaling supraspinal activation of large muscle groups.^49,50^ Interestingly, in subjects that showed high importance of decreased low-frequency activity during gait, GP beta power was upregulated during walking epochs, consistent with prior findings seen even in patients without significant gait abnormalities, which may indicate that this oscillatory change is not unique to the parkinsonian state.^51^ In contrast, for subjects where models showed high beta band importance, GP beta activity during walking was lower compared to the non-walking state, which may suggest a more pathological gait pattern that require greater beta desynchronization to promote an antikinetic state.^50–55^ The specificity of this finding, however, may be affected by the relationship between medication state and beta power, requiring further investigation. Overall, it is possible that the balance between beta desynchronization and low-frequency synchronization are necessary for gait – with the former releasing the ‘brake’ and the latter activating motion.

The ability to distinguish walking and non-walking states is critical to the treatment of advanced gait disorders in PD, which manifest as reduced gait speed,^25,56^ shorter step length,^56,57^ and increased interlimb asymmetry,^58,59^ amplifying the risk of falls and diminishing quality of life.^60–64^ Despite the profound consequences of impaired gait, the treatment of gait disturbances with conventional clinical stimulation has been a challenge. Early trials of adaptive DBS for PD have used cortical or subcortical control signals to selectively regulate stimulation amplitude during periods of symptom worsening, demonstrating improved symptom control.^23,24,33,34^ While pilot studies have produced nearly uniformly encouraging findings, adaptive approaches to gait dysfunction remain unexplored due to our limited understanding of neural changes in the real world. With the pipeline discussed in this paper, we propose one method of identifying cortical-pallidal biomarkers that can be used on-board embedded systems to automatically switch stimulation from conventional clinical settings when patients are at rest to gait-optimized frequencies when patients are walking.

Despite the ability to decode movement state in all hemispheres with high sensitivity and specificity, we observed significant variation in the cortical-pallidal biomarkers enabling this performance. Differences in movement state biomarkers may be explained by variance in gait dysfunction severity, PD phenotype, or electrode placement.^21^ Additionally, these biomarkers may be influenced by the amount of time spent in the various gait cycle phases by each subject as our approach averages across many individual cycles. Biomarkers also showed a range of sensitivity and specificity. In practical implementation, these biomarkers will likely be fine-tuned with threshold modification due to the tradeoffs in optimization. In other words, inappropriately switching to lower-frequency settings while a patient is at rest sacrifices appendicular symptom control and the opposite error may lead to substandard improvement in gait functions. To what extent either of these faults is tolerable may very well be patient- and impairment-dependent, which further emphasizes the value of a customizable pipeline for biomarker identification.

In this study, we also demonstrated the viability of an ambulatory rather than in-laboratory data collection pipeline. Advances in sensor technology have increasingly improved our ability to capture and study neural activity in naturalistic environments. Here, we illustrated the successful use wearable devices to accurately identify walking states in subjects with a wide range of pathologic gait patterns. This permitted the collection of large datasets across a range of days for all subjects at home more efficiently and dynamically than would have been possible under laboratory conditions.^5^ The ability to synchronize high-quality behavioral data with neural signals represents a major advancement in our ability to derive ecologically valid biomarkers for BCIs.^65^ Beyond research, this development may also hold value for clinical applications, including for remote outpatient therapy optimization and expansion of care to medically underserved areas.^66–68^

A primary limitation of our study is the small sample size given the invasive nature of surgical studies with investigation devices here. Future studies with larger cohorts and healthy controls are warranted to further elucidate supraspinal locomotor control mechanisms in neurotypical and pathological states. Another limitation of our study is the manual approach to neural and kinematic signal alignment, which is time-intensive and intractable for processing larger datasets. Considering the translational value of this work, the scalability of a naturalistic pipeline will be better addressed in the future by automatic/Bluetooth synchronization of external wearable devices with implantable neurostimulators.

Overall, the identification of effective gait state biomarkers is critical to a variety of diseases. While the pipeline discussed in this paper identifies oscillatory signatures will enable DBS devices to address both appendicular and axial dysfunction in PD, our approach is generalizable to the many neurological diseases impairing movement. Importantly, the pipeline discussed here offers a way to collect high-quality, longitudinal neural data in the real world. The insights gained from ecological data collection will advance therapy and accelerate BCIs across a multitude of debilitating conditions.

## METHODS

### Patient selection

Subjects with idiopathic PD were recruited from those being evaluated for DBS surgery at the University of California, San Francisco (2 male, 2 female, age range: 62-68 years, disease duration range: 4-21 years) **(Figure 2a)**. Inclusion criteria included the absence of significant cognitive impairment, ability to comply with study follow-up visits, Unified Parkinson’s Disease Rating Score between 20 to 80 and an improvement of at least 30% in the baseline on-medication score compared to the off-medication score, and gait impairments (slowed gait, shuffling steps, postural instability, or freezing of gait). Subjects were assessed and diagnosed with PD by neurologists specialized in movement disorders. All participants provided written informed consent. Institutional Review Board (IRB) at the University of California, San Francisco provided formal ethical approval for this study (IRB# 20-32847).

### DBS implantation and electrode reconstruction

The subjects enrolled in this study underwent surgical implantation of quadripolar DBS leads into the globus pallidus (Model 3389, Medtronic; contact length: 1.5 mm; intercontact distance: 2.0 mm) and subdural quadripolar paddles overlying the sensorimotor cortices (Model 0913025, Medtronic; contact diameter: 4 mm; intercontact distance: 10 mm). Two subjects were implanted unilaterally (left hemisphere) and two were implanted bilaterally. Electrodes were connected to investigational bidirectional implantable pulse generators placed in a superficial pocket over the ipsilateral pectoralis muscle (Summit RC+S Model B35300R, Medtronic). Further details of the surgical implantation procedure have been reported previously.^34,69,70^

Localization of depth and cortical electrodes was performed using advanced image processing pipelines (Lead-DBS, LeGUI) (**Figure 2b**).^71–74^ High-resolution postoperative CT images were coregistered to preoperative T1-weighted 3T MRI scans using a rigid, linear affine transformation. Coregistration accuracy was visually verified and refined when necessary using an additional brain shift correction routine to align subcortical anatomy. Subcortical electrode artifacts were identified on postoperative CT images and matched to known electrode geometry. Cortical electrodes were projected onto MRI-rendered pial surfaces and manually adjusted if needed.

### Summit RC+S device and neural recording preprocessing

The Summit RC+S device (Medtronic, model B35300R) is an investigational, rechargeable, bidirectional implantable neurostimulator (INS) that can stream four bipolar time domain electrode channels at a sampling rate of 500 Hz concurrent with delivery of therapeutic stimulation through a maximum of two quadripolar leads (**Figure 1a**). The INS also contains an embedded accelerometer which samples at 64 Hz. Further details of the Summit RC+S device can be found in previous publications.^69^

Data from the RC+S devices were extracted and analyzed using open-source code: https://github.com/openmind-consortium/Analysis-rcsdata. Neural signals from the GP, M1, and PM were bandpass filtered from 1 to 150 Hz using a 6^th^ order Butterworth filter. Signals were examined for any stimulation or ECG artifacts and template subtraction was used to identify and remove all instances of these artifacts: https://github.com/lhart1216/PerceptHammer. Epochs with RC+S packet loss or idiosyncratic artifacts were excluded from analysis.

### Rover wearable device

The Rover wearable device (WD) consists of a 38.1 mm x 51.2 mm x 13.7 mm, lightweight (53.86 g) rechargeable inertial sensor module paired with a fabric strap allowing the device to be worn around the ankle (**Figure 1b**). The 9-axis motion sensor module contains a triaxial gyroscope, triaxial accelerometer, and triaxial magnetometer which sample at 100 Hz and record data locally on a secure digital (SD) card. WD data were uploaded by subjects to the secure Rover cloud. Raw WD data were extracted and formatted with custom code.

Once a WD recording was uploaded to the cloud, the Rover Gait Analysis System (GAS) was used to generate a multiple-page gait analysis (MPGA) spreadsheet. Each MPGA summarized activity details (e.g., recording duration, total walking time, total distance walked, etc.) for each recording. In addition, it provided a list of time-stamped strides for both legs with gait cycle time (s), length (cm), swing period (s), and heading (°) recorded. MPGA reports were used to calculate average gait metrics for all subjects **(Supplementary Table S1)**.

### Validation of wearable device movement state identification

We performed independent validation of the Rover WD’s ability to accurately identify movement states in our cohort using kinematic data from observed walking and non-walking periods collected in a controlled environment. Patients were equipped with the Rover WD and Delsys sensors, which consist of two Avanti force-sensitive resistor (FSR) adapters, two Avanti goniometer adapters, and two Trigno surface electromyography (EMG) sensors with built-in accelerometers. The goniometer adapters were placed on the shanks bilaterally. EMG sensors were placed on top of the RC+S INS. FSR adapters were each attached to four FSRs (Delsys, model DC:F01) under the calcaneus, hallux, first metatarsal, and fifth metatarsal. Under supervision, subjects performed overground walking loops with interposing periods of standing, arm swing, or seated rest during which data were recorded from the RC+S, Rover WD, and Delsys devices. See **Supplementary Figure S1a** for sample trials for all subjects.

Following data collection, signals from these three devices were aligned with peak-to-peak matching across acceleration signals from the ipsilateral RC+S INS, Rover WD, and Delsys Trigno EMG accelerometer using a custom-built GUI in Matlab (Mathworks Inc., Natick, MA, United States, version 2021b). Gait cycles were identified using Delsys FSR data; individual cycles were defined as the time between two consecutive heel strikes (calcaneal FSR signal crossing 5% threshold in positive direction). Each recording of kinematic data was partitioned into 1-second epochs and each epoch was labeled as a walking period if it contained any portion of a gait cycle and non-walking if it contained no portions of any gait cycle. These labels were verified with review of video recordings taken of subjects’ activity during these sessions and manually adjusted if necessary.

To then compare Rover WD-based labeling of movement state with the ground-truth labels, all 1-second epochs were classified using the time-stamped stride lists from the Rover MPGA. Epochs composed of any left or right leg strides – determined by calculating the overlap of stride times and epoch times – were considered walking. Epochs with no overlapping left and right leg stride times were considered non-walking. WD-derived labels were then compared to ground-truth labels; accuracy, sensitivity, and specificity were calculated for each subject **(Supplementary Figure S1b)**.

### Neural and kinematic signal alignment

Subjects streamed neural data while wearing the Rover WD during normal activities of daily living several times weekly, as continuously as possible. Data from RC+S and WD devices for each subject were uploaded to a secure cloud folder accessible only by the research team. RC+S and WD recordings from the same days were identified.

A custom-built GUI in MATLAB was used to manually align x-axis acceleration signals from the RC+S and bilateral Rover WD by matching acceleration peak-to-peak across the devices. All alignments were reviewed extensively to ensure synchronization between RC+S and Rover acceleration signals.

### Movement state labeling

Each INS-WD alignment was partitioned into continuous 10-second epochs. The associated time-stamped MPGA stride lists and WD accelerometry signals were utilized to label each epoch with complete data as walking or non-walking. Epochs composed of greater than 50% (5 seconds) of left and right leg strides – determined by the overlap of stride times and epoch times – were considered walking. Epochs with no overlapping left and right leg stride times were labeled as non-walking. Remaining epochs were excluded from labeling and analysis as these periods were considered transition periods.

### Canonical frequency band analysis

Neural time-domain data from each 10-second epoch were converted to the frequency domain using Welch’s method (MATLAB ‘pwelch’ function; 1 second windows with 50% overlap). Average power was calculated within canonical frequency bands: delta (δ; 1-4 Hz), theta (θ; 4-8 Hz), alpha (ɑ; 8-13 Hz), beta (β; 13-30 Hz), and low gamma (low γ; 30-50 Hz). Two-sided Wilcoxon rank-sum tests were used to compare power within canonical frequency bands between walking and non-walking epochs.

To assess the relative importance of each canonical frequency band to within-subject discrimination of walking and non-walking epochs, logistic regression models were trained and tested within 10-fold cross validation (70/30 train/test split) using average power within canonical frequency bands from GP, M1, and PM signals (totaling 15 features per model). All features were standardized using z-scoring within each frequency band. AUC was calculated for models using class probabilities. For subjects with bilateral implants, separate models were generated for each hemisphere. Model coefficients were linearly rescaled from −1 to 1 and plotted on colored heat maps to visualize relative feature importance.

### Personalized gait biomarker identification

While canonical frequency bands offer the advantage of standardized comparison across individuals, they also carry several limitations including the potential masking of narrowband peaks.^35^ We therefore employed a data-driven approach to identify individualized frequency bands that may serve as potential endogenous biomarkers for movement state classification. To generate new candidate bands, we calculated average power within all bands of varying sizes from 1 Hz (e.g. [1-2 Hz], [2-3 Hz], etc.) to 49 Hz (i.e. [1-50 Hz]), thereby generating 1225 features per region.

Random forest (RF) models were used to identify the most important frequency bands as RF has been shown to overcome limitations of other feature selection algorithms including sensitivity to collinearity and high dimensionality.^75,76^ Feature importance was calculated for each frequency band as the mean decrease in impurity (MDI), a metric measuring the increase in subset homogeneity – or Gini impurity – when each variable is used for node splitting. For each node, the Gini impurity was calculated as follows, where *p_i_* is the proportion of samples belonging to class *c*:

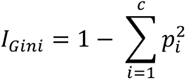

Mean feature importance was calculated from 1,000 RF iterations. We visualized feature importance for each hemisphere by rescaling the mean importance of each variable from 0 to 1 and plotting these values as overlapping bars for each frequency band.

We next turned to classification of movement state using these new frequency bands. Linear discriminant analysis (LDA) models were constructed to classify movement state for each subject using cortical-pallidal neural activity. LDA models were chosen as they generate feature weights that can be entered into the Summit RC+S on-board classifier when embedding adaptive architecture. Several types of LDA models were tested: single-region models using signals from one region (GP, M1, PM), multi-region models using signals from two regions (GP + M1, GP + PM, M1 + PM), and complete models using signals from all three regions (GP + M1 + PM).

Each LDA model was trained and tested with 10-fold cross validation; for each fold, the model was trained with 70% of the data and tested on the remaining 30%. Data were stratified to balance the representation of walking and non-walking epochs (50/50) in the training and testing sets. All features were standardized using z-scoring. Mean accuracy, AUC, sensitivity, specificity, and positive predictive value (PPV) were calculated.

### Simulation of RC+S on-board classification

The implementation of closed-loop aDBS architecture requires conformation to several practical constraints that differentiate it from the previous benchtop analyses. Notably, the RC+S device converts neural time-domain data to the frequency domain and calculates power features in a distinct manner, which has been discussed in previous publications.^77^ Furthermore, the LDA classifier on-board the RC+S device is limited to a maximum of four features with no more than two features from a single channel.

To best simulate on-board classification of movement state, we regenerated LDA models that more closely matched the above constraints. Epochs were shortened to 5 second intervals to mimic more rapid changes and features were recalculated with the same method used by the Summit RC+S device (see **Supplementary Methods** for detail). LDA models were also limited to the same number of maximum features (four) with the same channel constraints (maximum two features per channel); models with one to four features meeting these requirements were tested. RF feature importance values as discussed in an earlier section were used to determine the most important features for use in these system-constrained models.

First, 10-fold cross validated LDA models using recalculated features from 5-second epochs and meeting feature requirements were trained and tested using at-home recordings. To further validate our simulation of on-board classification, we next trained system-constrained models using at-home data and tested these models on recordings of subjects in observed settings. During these in-lab trials, subjects performed overground walking loops with interposing periods of standing or seated rest. Similar to the prior at-home analysis, models with one to four features meeting the on-board classifer requirements were tested. These simulations of continuous on-board classification were visualized using custom code: https://github.com/Weill-Neurohub-OPTiMaL.

### General Statistical Methods

#### Correction for multiple comparisons

In analyses with multiple comparisons (e.g., comparison of power within canonical frequency bands between movement states), we accounted for the false discovery rate (FDR) using the Benjamini-Hochberg procedure.^78^

#### Empirical p-value calculation

For each classification model tested, we characterized chance-level performance by generating 1,000 (n) permuted null models of randomly shuffled movement state labels and calculating AUC for each model. We then calculated the number of permuted AUCs greater than each LDA or logistic regression model AUC (k) and corrected this empirical one-sided p-value with the following adjustment similarly used in previous studies to avoid underestimation^13,79^:

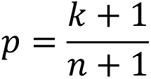

## Data Availability

Data from this study can be made available upon reasonable request, provided that patient confidentiality is maintained and disclosure standards are met.

## Notes

### Competing Interest Statement

The authors have declared no competing interest.

### Clinical Trial

NCT03582891

### Funding Statement

This research was supported by Michael J Fox Foundation (MJFF-010435) (DDW), NIH R01NS130183 (DDW), UCSF Catalyst Grant (DDW, RR, HFA), Tianqiao and Chrissy Chen Institute (DDW).

### Author Declarations

Institutional Review Board (IRB) at the University of California, San Francisco provided formal ethical approval for this study (IRB# 20-32847).

