## Supplementary Material for "At-Home Movement State Classification Using Totally Implantable Bidirectional Cortical-Basal Ganglia Neural Interface"

#### Supplementary Methods

**In-silico simulation of RC+S power feature calculation……………………………………………2**

**Supplementary References...……………………………………………………………………...3**

#### Supplementary Figures

**Figure S1. Validation of Rover wearable device movement state labeling….…………………4**

**Figure S2. Mean power spectral densities for walking and non-walking epochs…………..5**

#### Supplementary Tables

**Table S1. Summary of average gait parameters by subject……...……...….……..………..….6**

**Table S2. Summary of at-home neural and kinematic recordings.………..……...…….…….7**

**Table S3. Comparison of canonical band power between movement states………………..8**

**Table S4. Summary of logistic regression models for movement state classification…....9**

**Table S5. Most important feature from each region………………...…..…………………....10**

**Table S6. Summary of LDA classification of at-home movement state………...……..……11**

**Table S7. Summary of in-silico simulation of on-board classification…...…………..………12**

### In-silico simulation of RC+S power feature calculation

In order to simulate on-board classification of movement state, we performed offline calculation of signal power in the same way executed by the Summit RC+S device. This estimation has been discussed and compared to on-device output in previous publications, showing excellent replication.[1] First, the time-domain neural signals *s(n)* were converted from raw millivolt units to those of the device's internal system ($C_{RC+S}$: 48644.8683623726):

$$s(n)_{RC+S} = \left(s(n)_{mV} - \overline{s(n)_{mV}}\right) * \frac{\frac{250 * gain}{255} * C_{RC+S}}{1200}$$

Next, we calculated the overlap of these converted signals with a running Hann window (sampling rate: 500; FFT size: 256; actual FFT size: 250; default window load: 100%):

$$overlap = 1 - \left(\frac{500 * 256}{250}\right)$$

$$Hann\ window\ (n) = 0.5 * \left[1 - \cos\left(2\pi * \frac{n}{N}\right)\right], 0 \leq n \leq N$$

The single-sided FFT *S(k)* was then calculated for each window. Power features $P_{f1}^{f2}$ were calculated by summing power within all frequency bins within each defined frequency band:

$$P_{f1}^{f2} = \sum_{k=binA}^{binB} S(k)^2$$

**Figure S1. Validation of Rover wearable device movement state labeling.**

**a**

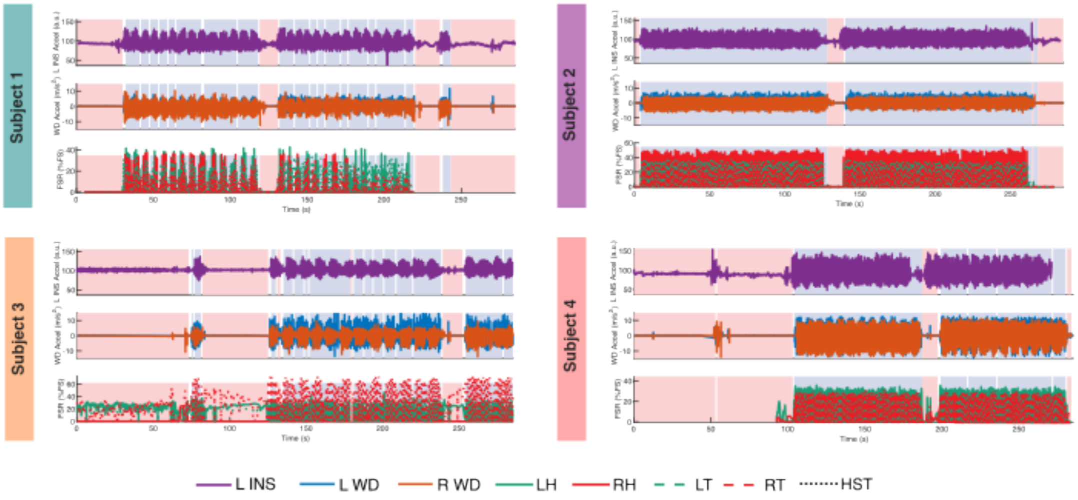


**b**

| Subject | Accuracy (%) | Sensitivity (%) | Specificity (%) |
|---|---|---|---|
| 1 | 95.8 | 98.9 | 94.7 |
| 2 | 99.0 | 94.4 | 100.0 |
| 3 | 97.3 | 95.6 | 99.1 |
| 4 | 96.6 | 95.0 | 100.0 |

**Legend. a.** Sample recordings of trials of observed walking and non-walking periods in controlled environments are shown for each subject. Signals from participants' INS (top), WD (middle), and force-sensitive resistors (FSRs) (bottom) placed under the calcaneus, hallux, first metatarsal (1MT), and fifth metatarsal (5MT) are shown. Epochs are shaded to indicate Rover WD-based labeling of movement state: blue (walking) and red (non-walking). **b.** Accuracy, sensitivity, and specificity of WD-based labeling is shown using FSR recordings with ground truth, verified with review of video recordings. 5MT = fifth metatarsal; FSR = force-sensitive resistor; HST = heel strike threshold; INS = implantable neurostimulator; LH = left heel; LT = left toe; RH = right heel; RT = right toe; Sig. = significance; WD = wearable device.

**Figure S2. Mean power spectral densities for walking and non-walking epochs.**

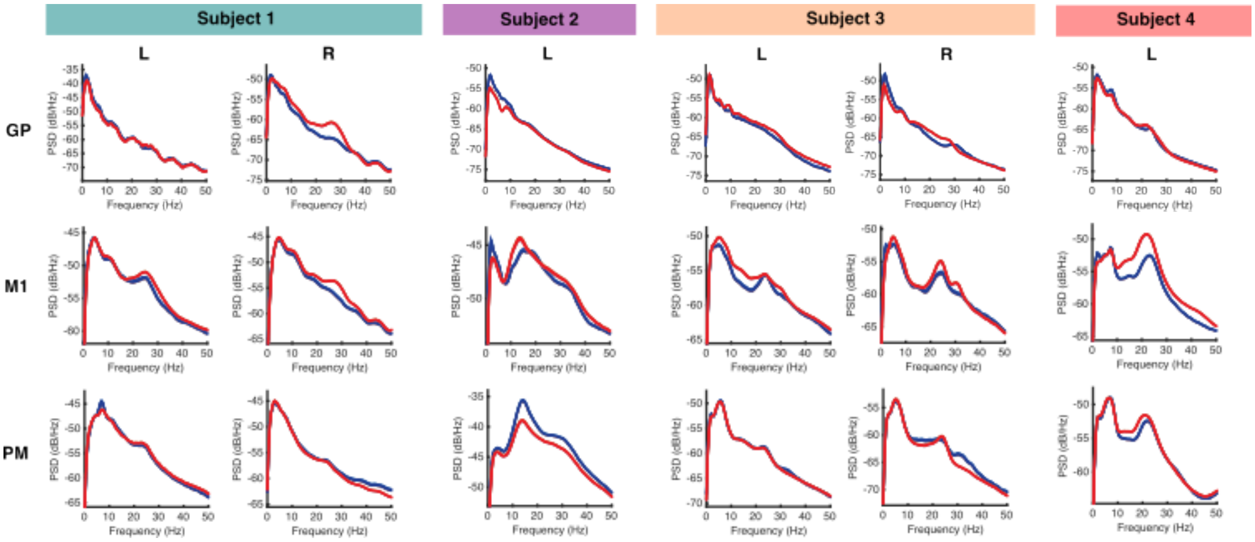


**Legend.** Welch's method was used to convert neural time-domain signals from GP, M1, and PM to the frequency domain. Mean power spectral densities are shown from 0 to 50 Hz for all walking (blue) and non-walking (red) 10-second epochs analyzed from each hemisphere. GP = globus pallidus; M1 = primary motor cortex; PM = premotor cortex.

**Table S1. Summary of average gait parameters by subject.**

| Subject | Average Gait Metric | | | | | | | | |
|---|---|---|---|---|---|---|---|---|---|
| | Stride Length (cm) | Stride Velocity (cm) | Cadence (steps/min) | Gait Cycle Time (s) | Stance Time (s) | Swing Time (s) | Stance Period (%GCT) | Swing Period (%GCT) | Swing Velocity (cm/s) |
| **1** | 85.6 | 66.4 | 93.9 | 1.3 | 1.0 | 0.3 | 76.1 | 23.9 | 293.5 |
| **2** | 140.5 | 118.2 | 100.8 | 1.2 | 0.8 | 0.4 | 66.9 | 33.1 | 362.4 |
| **3** | 96.4 | 75.4 | 98.7 | 1.3 | 1.0 | 0.3 | 73.6 | 26.4 | 306.2 |
| **4** | 89.8 | 87.0 | 116.8 | 1.0 | 0.8 | 0.3 | 75.1 | 24.9 | 361.6 |

**Legend.** Average stride length, stride velocity, cadence, gait cycle time, stance time, swing time, stance period, swing period, and swing velocity are shown for all subjects using the Rover WD GAS. GAS = gait analysis system; GCT = gait cycle time; WD = wearable device.

**Table S2. Summary of at-home neural and kinematic recordings.**

| Subject | Total Analyzed Recording Duration (s) | Number of Recording Days (d) |
|---|---|---|
| **1** | 118,660 | 7 |
| **2** | 33,820 | 17 |
| **3** | 59,860 | 5 |
| **4** | 91,900 | 23 |

**Legend.** Total analyzed recording duration and the number of individual days on which recordings were performed are shown for all walking and non-walking periods extracted from at-home neural and kinematic recordings for each subject.

### Table S3. Comparison of canonical band power between movement states.

| Subject | Region | Median Power within Canonical Frequency Band (dB/Hz) | | | | | | | | | | | | | | |
|---|---|---|---|---|---|---|---|---|---|---|---|---|---|---|---|---|
| | | δ (1-4 Hz) | | | Θ (4-8 Hz) | | | α (8-13 Hz) | | | β (13-30 Hz) | | | $\gamma_{low}$ (30-50 Hz) | | |
| | | NW | W | p | NW | W | p | NW | W | p | NW | W | p | NW | W | p |
| 1 | L GP | -40.9 | -40.0 | 1.3e-22 | -48.5 | -47.5 | 6.0e-44 | -54.2 | -53.4 | 3.1e-20 | -60.8 | -60.6 | 0.62 | -69.5 | -68.8 | 3.5e-25 |
| | L M1 | -47.3 | -47.0 | 8.2e-8 | -46.6 | -46.8 | 0.033 | **-48.9** | **-49.4** | **6.7e-5** | **-51.2** | **-52.0** | **1.3e-6** | -57.3 | -58.2 | 6.9e-24 |
| | L PM | -49.3 | -49.4 | 0.089 | -48.9 | -45.7 | 3.8e-30 | -47.9 | -47.7 | 0.016 | -52.8 | -52.9 | 0.50 | -60.9 | -61.1 | 1.4e-6 |
| | R GP | -50.6 | -50.5 | 0.37 | -51.8 | -53.3 | 7.1e-43 | -55.8 | -56.9 | 1.4e-52 | -61.4 | -63.3 | 7.8e-75 | -69.5 | -70.1 | 5.4e-22 |
| | R M1 | -47.3 | -48.0 | 2.3e-4 | -46.2 | -47.0 | 7.8e-10 | **-47.9** | **-48.9** | **9.8e-8** | **-51.5** | **-53.8** | **3.6e-18** | -59.0 | -60.8 | 2.2e-30 |
| | R PM | -45.6 | -46.0 | 1.1e-9 | -46.9 | -46.9 | 0.12 | -50.6 | -50.5 | 0.012 | -56.1 | -56.0 | 3.7e-4 | -62.2 | -61.7 | 2.0e-21 |
| 2 | L GP | -56.9 | -53.5 | 1.3e-153 | -60.1 | -57.1 | 2.5e-161 | -61.3 | -59.8 | 6.9e-81 | -66.4 | -66.0 | 6.0e-21 | -73.1 | -72.8 | 7.5e-57 |
| | L M1 | -47.0 | -45.5 | 1.6e-57 | -48.1 | -47.5 | 1.7e-8 | **-47.0** | **-47.7** | **9.2e-6** | **-45.8** | **-46.5** | **5.0e-6** | -50.4 | -51.2 | 2.6e-20 |
| | L PM | -45.3 | -44.4 | 3.4e-19 | -44.7 | -43.6 | 2.4e-15 | -41.6 | -40.0 | 5.0e-32 | -40.7 | -38.9 | 3.8e-79 | -47.6 | -46.4 | 5.8e-100 |
| 3 | L GP | -52.2 | -52.2 | 0.65 | -56.2 | -56.6 | 2.2e-16 | -57.9 | -58.9 | 1.6e-51 | -61.4 | -62.4 | 1.6e-43 | -70.3 | -70.9 | 1.2e-40 |
| | L M1 | -51.6 | -52.1 | 1.2e-9 | -50.8 | -51.9 | 1.3e-55 | **-53.8** | **-55.0** | **2.8e-31** | **-56.2** | **-57.2** | **1.3e-15** | -61.1 | -61.5 | 1.5e-10 |
| | L PM | -52.4 | -52.2 | 7.7e-3 | -50.7 | -50.5 | 0.13 | -55.8 | -55.8 | 0.74 | -59.6 | -59.3 | 0.014 | -66.4 | -66.3 | 8.9e-4 |
| | R GP | -53.6 | -50.3 | 1.4e-87 | -57.6 | -55.6 | 4.4e-65 | -59.5 | -59.2 | 3.6e-4 | -63.8 | -65.0 | 2.6e-38 | -71.4 | -70.7 | 4.8e-13 |
| | R M1 | -53.7 | -53.2 | 2.2e-10 | -52.1 | -53.3 | 1.0e-18 | **-56.5** | **-57.0** | **5.9e-7** | **-57.7** | **-58.6** | **1.2e-9** | -62.8 | -62.9 | 0.72 |
| | R PM | -55.7 | -56.1 | 0.036 | -54.5 | -54.7 | 0.027 | -59.8 | -59.5 | 1.3e-3 | -62.2 | -61.6 | 1.3e-8 | -68.3 | -67.4 | 1.8e-58 |
| 4 | L GP | -53.5 | -52.9 | 1.8e-65 | -56.3 | -55.6 | 2.5e-77 | -59.3 | -58.9 | 7.7e-28 | -64.7 | -65.2 | 1.0e-29 | -72.9 | -72.7 | 1.6e-67 |
| | L M1 | -52.5 | -52.1 | 6.7e-54 | -52.4 | -51.8 | 3.3e-6 | **-53.8** | **-54.6** | **5.6e-48** | **-51.9** | **-54.9** | **4.6e-263** | -60.6 | -61.9 | 3.2e-163 |
| | L PM | -51.5 | -51.7 | 2.2e-6 | -49.4 | -49.5 | 0.077 | -52.7 | -52.9 | 2.9e-9 | -53.8 | -54.7 | 9.9e-56 | -61.7 | -61.8 | 2.6e-3 |

**Legend.** Median power is shown across five canonical frequency bands of interest: delta (δ), theta (Θ), alpha (α), beta (β), and low gamma ($\gamma_{low}$) for GP, M1, and PM signals. Power differences between walking (W) and non-walking (NW) epochs were assessed using two-sided Wilcoxon rank sum tests with Benjamini-Hochberg correction for multiple comparisons. Uncorrected p-values are shown. Boxes are shaded to indicated bands where power was significantly higher during walking (blue), significantly higher during non-walking (red), or not significantly different between movement states (gray). Alpha and beta power in M1 was higher during non-walking epochs in all subjects, indicated by hatched boxes. GP = globus pallidus; M1 = primary motor cortex; PM = premotor cortex.

### Table S4. Summary of logistic regression models for movement state classification.

| Subject | Side | Rescaled Coefficients | | | | | | | | | | | | | | | AUC | Sig. (p) |
|---|---|---|---|---|---|---|---|---|---|---|---|---|---|---|---|---|---|---|
| | | GP | | | | | M1 | | | | | PM | | | | | | |
| | | δ | Θ | α | β | $\gamma_{low}$ | δ | Θ | α | β | $\gamma_{low}$ | δ | Θ | α | β | $\gamma_{low}$ | | |
| 1 | L | -0.015 | 0.33 | 0.68 | -1.0 | 1.0 | 0.39 | -0.22 | 0.044 | 0.49 | -0.40 | -0.64 | 0.78 | -0.10 | 0.44 | -0.38 | 0.79 | <0.001 |
| | R | 0.73 | -0.68 | 0.25 | -0.77 | 0.28 | 0.35 | -0.18 | 0.12 | 1.0 | -1.0 | -0.26 | 0.065 | 0.12 | 0.12 | -0.064 | 0.87 | <0.001 |
| 2 | L | 0.33 | 0.65 | 0.085 | -0.76 | 0.45 | 0.76 | 0.47 | -1.0 | 0.18 | -0.39 | 0.44 | -0.10 | 1.0 | 0.44 | 0.45 | 0.94 | <0.001 |
| 3 | L | 0.28 | -0.23 | -0.42 | -0.057 | -0.60 | -0.015 | -1.0 | -0.30 | 0.18 | 0.72 | 0.15 | 0.31 | -0.32 | 1.0 | -0.30 | 0.81 | <0.001 |
| | R | 0.89 | 1.0 | -0.15 | -1.0 | -0.19 | 0.28 | -0.37 | -0.21 | 0.41 | -0.89 | -0.39 | 0.14 | -0.18 | 0.18 | 0.98 | 0.93 | <0.001 |
| 4 | L | 0.10 | 0.46 | 0.33 | -0.32 | 0.21 | 0.99 | 1.0 | -0.076 | -0.32 | -1.0 | -0.29 | -0.16 | -0.21 | 0.89 | 0.53 | 0.93 | <0.001 |

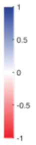


**Legend.** Logistic regression models were trained to classify movement state (walking vs. non-walking) using average power within canonical frequency bands from GP, M1, and PM signals, separately for each hemisphere. Model coefficients are shown, rescaled from -1 to 1. Mean AUC is reported for each model, with 10-fold cross validation. Empirical one-sided p-values were calculated by randomly shuffling true labels 1,000 times and determining the AUC for each permutation. GP = globus pallidus; M1 = primary motor cortex; PM = premotor cortex; Sig. = significance.

**Table S5. Most important feature from each region.**

| Subject | Side | GP | | M1 | | PM | |
|---|---|---|---|---|---|---|---|
| | | Band (Hz) | Importance | Band (Hz) | Importance | Band (Hz) | Importance |
| 1 | L | 6-7 | 1.0 | 1-2 | 0.69 | 6-7 | 0.62 |
| | R | 27-29 | 1.0 | 1-4 | 0.49 | 2-3 | 0.16 |
| 2 | L | 1-6 | 1.0 | 2-7 | 0.50 | 14-18 | 0.03 |
| 3 | L | 5-6 | 0.96 | 4-8 | 1.0 | 32-36 | 0.35 |
| | R | 2-6 | 1.0 | 27-32 | 0.06 | 29-36 | 0.31 |
| 4 | L | 1-10 | 0.29 | 18-23 | 1.0 | 20-24 | 0.22 |

**Legend.** Random forest models were trained to classify movement state (walking vs. non-walking) using average power within all possible frequency bands of sizes 1 to 49 Hz (within the 1-50 Hz range of interest) from GP, M1, and PM signals, separately for each hemisphere. Feature importance was determined by calculating the mean decrease in impurity for each frequency band. The most important frequency band in each region is shown for each hemisphere, along with its normalized importance value. GP = globus pallidus; M1 = primary motor cortex; PM = premotor cortex.

### Table S6. Summary of LDA classification of at-home movement state.

| Subject | Side | Model | AUC | Sensitivity (%) | Specificity (%) | PPV (%) | Sig. (p) |
|---|---|---|---|---|---|---|---|
| 1 | L | **Single-Region** | | | | | |
| | | GP | 0.76 ± 0.02 | 79.6 ± 7.3 | 63.4 ± 7.5 | 69.0 ± 2.7 | <0.001 |
| | | M1 | 0.67 ± 0.02 | 71.5 ± 8.8 | 56.3 ± 7.9 | 62.5 ± 2.8 | <0.001 |
| | | PM | 0.70 ± 0.02 | 65.9 ± 8.2 | 68.0 ± 8.7 | 68.2 ± 4.6 | <0.001 |
| | | **Multi-Region** | | | | | |
| | | GP + M1 | **0.77 ± 0.02** | **74.8 ± 4.9** | **69.7 ± 6.5** | **71.9 ± 4.7** | <0.001 |
| | | M1 + PM | 0.73 ± 0.02 | 73.9 ± 9.3 | 64.5 ± 8.2 | 68.5 ± 4.1 | <0.001 |
| | | GP + PM | **0.77 ± 0.02** | **77.5 ± 5.2** | **70.1 ± 6.0** | **72.4 ± 4.1** | **<0.001** |
| | | **Complete** | | | | | |
| | | GP + M1 + PM | **0.77 ± 0.02** | **76.4 ± 7.9** | **66.9 ± 7.3** | **69.7 ± 3.9** | **<0.001** |
| | R | **Single-Region** | | | | | |
| | | GP | **0.86 ± 0.02** | **85.6 ± 5.5** | **73.1 ± 5.6** | **75.3 ± 2.9** | **<0.001** |
| | | M1 | 0.83 ± 0.02 | 85.0 ± 6.3 | 68.6 ± 7.4 | 72.6 ± 4.6 | <0.001 |
| | | PM | 0.73 ± 0.02 | 72.2 ± 8.3 | 64.6 ± 8.4 | 68.5 ± 2.6 | <0.001 |
| | | **Multi-Region** | | | | | |
| | | GP + M1 | 0.84 ± 0.02 | 83.9 ± 7.1 | 71.2 ± 8.0 | 74.8 ± 3.9 | <0.001 |
| | | M1 + PM | 0.82 ± 0.02 | 81.4 ± 6.8 | 69.2 ± 6.1 | 73.2 ± 2.9 | <0.001 |
| | | GP + PM | 0.84 ± 0.01 | 81.7 ± 4.1 | 73.2 ± 3.2 | 75.8 ± 2.6 | <0.001 |
| | | **Complete** | | | | | |
| | | GP + M1 + PM | 0.83 ± 0.02 | 78.6 ± 3.1 | 75.1 ± 4.6 | 76.7 ± 3.0 | <0.001 |
| 2 | L | **Single-Region** | | | | | |
| | | GP | 0.89 ± 0.01 | 86.2 ± 3.1 | 78.4 ± 3.5 | 80.6 ± 2.2 | <0.001 |
| | | M1 | 0.85 ± 0.01 | 81.2 ± 3.5 | 73.8 ± 5.1 | 75.5 ± 3.4 | <0.001 |
| | | PM | 0.78 ± 0.02 | 75.1 ± 6.7 | 70.2 ± 5.4 | 71.7 ± 2.2 | <0.001 |
| | | **Multi-Region** | | | | | |
| | | GP + M1 | 0.92 ± 0.01 | 87.2 ± 3.5 | 82.5 ± 4.5 | 83.3 ± 3.2 | <0.001 |
| | | M1 + PM | 0.91 ± 0.01 | 88.1 ± 2.5 | 81.3 ± 3.3 | 82.5 ± 3.5 | <0.001 |
| | | GP + PM | 0.92 ± 0.01 | 90.5 ± 2.3 | 83.5 ± 2.3 | 84.2 ± 2.1 | <0.001 |
| | | **Complete** | | | | | |
| | | GP + M1 + PM | **0.94 ± 0.01** | **91.6 ± 2.0** | **86.6 ± 2.1** | **87.4 ± 1.7** | <0.001 |
| 3 | L | **Single-Region** | | | | | |
| | | GP | **0.91 ± 0.02** | **89.0 ± 4.1** | **82.1 ± 4.7** | **84.3 ± 3.3** | **<0.001** |
| | | M1 | 0.76 ± 0.02 | 70.6 ± 6.3 | 71.3 ± 7.4 | 70.2 ± 3.3 | <0.001 |
| | | PM | 0.68 ± 0.02 | 67.3 ± 12.9 | 63.4 ± 13.6 | 65.0 ± 4.5 | <0.001 |
| | | **Multi-Region** | | | | | |
| | | GP + M1 | **0.91 ± 0.01** | **81.8 ± 4.6** | **86.7 ± 3.7** | **86.5 ± 2.9** | **<0.001** |
| | | M1 + PM | 0.78 ± 0.02 | 72.5 ± 7.1 | 73.1 ± 6.8 | 73.6 ± 5.2 | <0.001 |
| | | GP + PM | 0.90 ± 0.02 | 85.1 ± 3.3 | 83.6 ± 5.1 | 84.1 ± 4.0 | <0.001 |
| | | **Complete** | | | | | |
| | | GP + M1 + PM | 0.89 ± 0.02 | 81.6 ± 2.8 | 83.3 ± 5.4 | 83.0 ± 5.0 | <0.001 |
| | R | **Single-Region** | | | | | |
| | | GP | **0.98 ± 0.01** | **95.8 ± 2.1** | **94.0 ± 2.9** | **94.1 ± 2.9** | **<0.001** |
| | | M1 | 0.87 ± 0.03 | 81.1 ± 8.3 | 83.2 ± 6.2 | 83.4 ± 4.8 | <0.001 |
| | | PM | 0.87 ± 0.03 | 80.8 ± 7.5 | 80.7 ± 9.0 | 81.0 ± 9.0 | <0.001 |
| | | **Multi-Region** | | | | | |
| | | GP + M1 | 0.93 ± 0.02 | 90.5 ± 6.0 | 84.8 ± 7.4 | 85.5 ± 5.6 | <0.001 |
| | | M1 + PM | 0.83 ± 0.04 | 72.0 ± 12.1 | 82.8 ± 5.8 | 80.0 ± 4.6 | <0.001 |
| | | GP + PM | 0.93 ± 0.02 | 89.9 ± 4.7 | 85.7 ± 4.0 | 86.2 ± 3.7 | <0.001 |
| | | **Complete** | | | | | |
| | | GP + M1 + PM | 0.74 ± 0.05 | 72.7 ± 6.6 | 70.9 ± 8.8 | 72.3 ± 7.0 | <0.001 |
| 4 | L | **Single-Region** | | | | | |
| | | GP | 0.87 ± 0.01 | 80.0 ± 2.9 | 78.9 ± 3.8 | 79.4 ± 2.6 | <0.001 |
| | | M1 | 0.90 ± 0.01 | 82.5 ± 2.7 | 83.3 ± 3.1 | 83.5 ± 2.3 | <0.001 |
| | | PM | 0.77 ± 0.01 | 75.0 ± 3.9 | 66.0 ± 4.0 | 68.9 ± 2.1 | <0.001 |
| | | **Multi-Region** | | | | | |
| | | GP + M1 | 0.94 ± 0.001 | 89.5 ± 2.4 | 85.5 ± 2.0 | 86.1 ± 1.3 | <0.001 |
| | | M1 + PM | 0.94 ± 0.01 | 86.7 ± 1.8 | 87.0 ± 2.6 | 87.1 ± 1.9 | <0.001 |
| | | GP + PM | 0.90 ± 0.01 | 82.5 ± 3.4 | 82.8 ± 2.5 | 83.2 ± 2.1 | <0.001 |
| | | **Complete** | | | | | |
| | | GP + M1 + PM | **0.96 ± 0.001** | **90.7 ± 1.8** | **89.5 ± 2.5** | **89.6 ± 2.3** | **<0.001** |

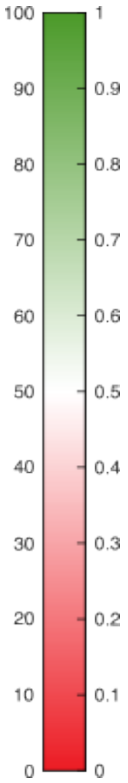


**Legend.** LDA models were trained to classify movement state (walking vs. non-walking) using average power within all possible frequency bands of sizes 1 to 49 Hz (within the 1-50 Hz range of interest) from combinations of GP, M1, and PM signals, separately for each hemisphere. Average AUC, sensitivity, specificity, and PPV are shown for each model using 10-fold cross-validation (Mean ± SD). Empirical one-sided p-values were calculated by randomly shuffling true labels 1,000 times and determining the AUC for each permutation. GP = globus pallidus; M1 = primary motor cortex; PM = premotor cortex; PPV = positive predictive value; SD = standard deviation.

### Table S7. Summary of in-silico simulation of on-board classification.

| Subject | Side | Number of Features | Train At-Home/Test At-Home | | | | | Train At-Home/Test In-Clinic | | | | |
|---|---|---|---|---|---|---|---|---|---|---|---|---|
| | | | AUC | Sensitivity (%) | Specificity (%) | PPV (%) | Sig. (p) | AUC | Sensitivity (%) | Specificity (%) | PPV (%) | Sig. (p) |
| 1 | L | 1 | 0.57 ± 0.01 | 60.3 ± 14.2 | 50.3 ± 14.9 | 55.4 ± 2.2 | 0.006 | 0.51 ± 0.03 | 54.3 ± 21.8 | 55.0 ± 22.6 | 56.4 ± 5.3 | 0.458 |
| | | 2 | 0.62 ± 0.01 | 51.1 ± 8.6 | 67.4 ± 8.5 | 61.3 ± 1.7 | <0.001 | 0.52 ± 0.05 | 55.7 ± 29.7 | 54.3 ± 30.5 | 58.7 ± 10.1 | 0.399 |
| | | 3 | 0.63 ± 0.01 | 59.6 ± 10.6 | 59.5 ± 10.3 | 59.9 ± 2.2 | <0.001 | 0.51 ± 0.02 | 47.2 ± 20.9 | 65.7 ± 22.0 | 61.6 ± 9.2 | 0.429 |
| | | 4 | 0.64 ± 0.01 | 71.0 ± 4.0 | 51.6 ± 4.1 | 59.5 ± 0.9 | <0.001 | 0.63 ± 0.04 | 74.3 ± 5.0 | 52.8 ± 8.4 | 61.5 ± 3.5 | 0.017 |
| | R | 1 | 0.71 ± 0.01 | 73.9 ± 0.7 | 62.5 ± 1.3 | 66.4 ± 0.6 | <0.001 | 0.86 ± 0.04 | 90.7 ± 1.3 | 78.4 ± 6.2 | 81.1 ± 4.4 | <0.001 |
| | | 2 | 0.71 ± 0.01 | 73.7 ± 0.6 | 62.3 ± 1.5 | 66.2 ± 0.8 | <0.001 | 0.87 ± 0.02 | 89.3 ± 2.4 | 81.3 ± 2.8 | 82.8 ± 2.1 | <0.001 |
| | | 3 | 0.70 ± 0.01 | 73.5 ± 0.8 | 62.4 ± 1.9 | 66.2 ± 1.2 | <0.001 | 0.51 ± 0.03 | 75.3 ± 38.1 | 30.9 ± 35.6 | 52.3 ± 1.3 | 0.391 |
| | | 4 | 0.75 ± 0.01 | 68.6 ± 2.2 | 68.8 ± 1.8 | 68.8 ± 0.8 | <0.001 | 0.50 ± 0.04 | 81.6 ± 28.2 | 25.3 ± 27.8 | 52.4 ± 2.1 | 0.505 |
| 2 | L | 1 | 0.42 ± 0.06 | 92.9 ± 4.0 | 16.0 ± 9.1 | 52.7 ± 2.0 | 1.000 | 0.50 ± 0.41 | 47.2 ± 0.2 | 87.0 ± 1.4 | 79.0 ± 0.8 | 0.477 |
| | | 2 | 0.62 ± 0.01 | 41.4 ± 2.7 | 75.8 ± 1.5 | 63.1 ± 0.7 | <0.001 | 0.80 ± 0.01 | 88.6 ± 0.3 | 71.1 ± 1.5 | 75.5 ± 0.9 | <0.001 |
| | | 3 | 0.75 ± 0.01 | 71.9 ± 2.7 | 68.1 ± 2.5 | 69.3 ± 1.0 | <0.001 | 0.74 ± 0.01 | 90.3 ± 1.5 | 60.0 ± 1.6 | 69.3 ± 0.7 | <0.001 |
| | | 4 | 0.75 ± 0.01 | 75.0 ± 1.7 | 67.1 ± 1.7 | 68.8 ± 0.7 | <0.001 | 0.75 ± 0.01 | 86.7 ± 2.5 | 61.5 ± 3.5 | 69.3 ± 1.5 | <0.001 |
| 3 | L | 1 | 0.72 ± 0.01 | 69.6 ± 7.7 | 62.6 ± 7.0 | 65.3 ± 2.0 | <0.001 | 0.77 ± 0.02 | 85.2 ± 0.9 | 67.8 ± 3.9 | 72.7 ± 2.3 | <0.001 |
| | | 2 | 0.68 ± 0.05 | 60.2 ± 4.8 | 67.8 ± 9.8 | 65.8 ± 4.4 | <0.001 | 0.70 ± 0.06 | 88.5 ± 2.0 | 55.9 ± 7.5 | 66.9 ± 3.1 | <0.001 |
| | | 3 | 0.72 ± 0.03 | 73.0 ± 3.8 | 60.1 ± 5.5 | 64.8 ± 2.3 | <0.001 | 0.78 ± 0.04 | 84.3 ± 8.2 | 65.4 ± 10.6 | 71.6 ± 5.6 | <0.001 |
| | | 4 | 0.74 ± 0.02 | 66.0 ± 6.8 | 67.1 ± 5.6 | 66.9 ± 2.2 | <0.001 | 0.78 ± 0.02 | 90.2 ± 7.5 | 62.0 ± 4.2 | 69.0 ± 2.0 | <0.001 |
| | R | 1 | 0.73 ± 0.01 | 67.4 ± 0.5 | 73.1 ± 1.2 | 71.5 ± 0.9 | <0.001 | 0.64 ± 0.02 | 57.0 ± 16.0 | 66.3 ± 13.9 | 64.4 ± 4.8 | 0.009 |
| | | 2 | 0.65 ± 0.03 | 71.0 ± 4.2 | 56.7 ± 8.1 | 62.4 ± 3.2 | <0.001 | 0.64 ± 0.02 | 53.0 ± 3.6 | 74.2 ± 6.2 | 67.7 ± 5.0 | 0.007 |
| | | 3 | 0.67 ± 0.01 | 64.7 ± 2.4 | 61.4 ± 2.7 | 62.6 ± 0.9 | <0.001 | 0.66 ± 0.02 | 61.7 ± 8.4 | 70.3 ± 7.8 | 68.0 ± 3.3 | 0.005 |
| | | 4 | 0.81 ± 0.01 | 76.2 ± 3.9 | 69.1 ± 4.6 | 71.3 ± 1.9 | <0.001 | 0.80 ± 0.02 | 73.0 ± 6.4 | 78.8 ± 6.8 | 78.1 ± 5.0 | <0.001 |
| 4 | L | 1 | 0.70 ± 0.00 | 87.5 ± 1.5 | 53.1 ± 1.7 | 65.1 ± 0.5 | <0.001 | 0.77 ± 0.01 | 87.2 ± 1.6 | 59.1 ± 1.8 | 68.1 ± 0.8 | <0.001 |
| | | 2 | 0.70 ± 0.01 | 87.7 ± 1.7 | 53.2 ± 1.7 | 65.2 ± 0.4 | <0.001 | 0.77 ± 0.01 | 86.7 ± 1.2 | 59.1 ± 1.6 | 67.9 ± 0.8 | <0.001 |
| | | 3 | 0.79 ± 0.01 | 83.3 ± 1.7 | 59.9 ± 1.3 | 67.5 ± 0.3 | <0.001 | 0.63 ± 0.01 | 85.7 ± 1.1 | 43.8 ± 1.5 | 60.4 ± 0.5 | <0.001 |
| | | 4 | 0.79 ± 0.01 | 81.4 ± 4.1 | 61.5 ± 4.2 | 68.0 ± 1.4 | <0.001 | 0.62 ± 0.02 | 88.8 ± 3.3 | 39.3 ± 4.3 | 59.4 ± 1.0 | <0.001 |

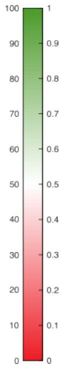


**Legend.** Performance of simulated on-board LDA classifiers are shown for models trained and tested on at-home data and models trained on at-home data and tested on in-clinic observed trials of overground walking with interspersed periods of standing or seated rest. Models with up to four features permissible for use in the on-board classifier were tested. Empirical one-sided p-values were calculated by comparing model performance with a permuted chance-level distribution (n = 1,000). LDA = linear discriminant analysis; PPV = positive predictive values; Sig. = significance.
